# A Hybrid Deep Learning-Mechanistic Modeling Framework for Dengue Transmission Dynamics in Guangdong, China

**DOI:** 10.1101/2025.10.04.25337267

**Authors:** Pei Yuan, Ning Wang, Qiyong Liu, Huaiping Zhu

**Affiliations:** School of Mathematical Sciences, Shenzhen University, 518061, China; Centre for Diseases Modelling (CDM) and Laboratory of Mathematical Parallel Systems (LAMPS), York University, Toronto, ON, M3J1P3, Canada; National Key Laboratory of Intelligent Tracking and Forecasting for Infectious Diseases, National Institute for Communicable Disease Control and Prevention, Chinese Center for Disease Control and Prevention; WHO Collaborating Centre for Vector Surveillance and Management, Beijing 102206, China; Department of Mathematics and Statistics, York University, Toronto, ON, M3J1P3, Canada

**Keywords:** Dengue fever, Climate factors, Monitoring data, Hybrid modelling, SIR model, RNN, Long short-term memory, simulations

## Abstract

Dengue fever is a mosquito-borne viral disease with strong seasonality, periodicity, and spatial heterogeneity, posing a persistent global public health threat. We present a hybrid modeling framework that couples a recurrent neural network (RNN) with a typical Susceptible–Infected–Recovered (SIR) compartmental model to investigate dengue transmission dynamics in Guangdong, China, from 2016 to 2024. By incorporating mosquito surveillance data, meteorological variables (temperature, humidity, and precipitation), public health intervention intensity, and reported dengue cases, our model captures the complex interactions among climate, vector density, intervention policies, and disease spread. After evaluating multiple RNN architectures, the Long Short-Term Memory (LSTM) model was selected for its superior performance in predicting the mosquito ovitrap index (MOI) from climatic variables, providing a data-driven proxy for vector abundance used in the SIR model. The hybrid model is constructed upon a partially observed Markov process (POMP) and calibrated using iterated filtering for parameter optimization. Model results identify climate variables as dominant drivers of dengue transmission, while non-pharmaceutical interventions (NPIs) during the COVID-19 pandemic significantly suppressed case numbers. Scenario analysis indicates that moderate NPIs could effectively reduce outbreak magnitude, with the model estimating that approximately 46,120 dengue cases were averted during the pandemic period. This study highlights the utility of integrating deep learning with mechanistic modeling to improve understanding and forecasting of vector-borne disease dynamics. The proposed framework offers a robust and interpretable approach for developing climate- and vector-informed early warning systems and informing data-driven public health decision-making.

**Author Summary:** Dengue fever is a mosquito-borne disease that causes large outbreaks in many tropical and subtropical regions, including southern China. Predicting when and where outbreaks will occur is difficult because dengue transmission is influenced by many factors, such as climate, mosquito populations, and public health measures. In this study, we developed a hybrid modeling framework that integrates artificial intelligence with mechanistic disease modeling. Specifically, we used a deep learning method to predict mosquito abundance from climate conditions and then linked to a mathematical model for dengue transmission. Applying this approach to data from Guangdong Province, China between 2016 and 2024, we found that climate strongly influences mosquito biting behavior, which in turn drives dengue transmission. We also showed that non-pharmaceutical interventions during the COVID-19 pandemic substantially reduced dengue cases, with our model estimating that more than 46,000 infections were prevented. Our innovative approach demonstrates how combining modern machine learning with mechanistic models can improve our ability to understand and forecast outbreaks of mosquito-borne diseases. Our framework may help inform early warning systems and guide public health strategies to reduce the burden of dengue and other vector-borne diseases.

## 1. Introduction

Dengue fever is one of the fastest spreading vector-borne diseases globally [1], with widespread endemicity in tropical and subtropical regions such as Africa, the Americas, Southeast Asia, and the Western Pacific. Nearly half of the world’s population is at risk of dengue infection [2]. In recent years, the geographic range of dengue has expanded northward due to accelerating climate change and intensified human mobility [3,4]. The global dengue burden has risen dramatically, reaching an unprecedented peak in 2024, marking the worst year on record [4], with over 7.6 million confirmed cases and more than 10,000 deaths reported worldwide [5]. In China, Guangdong Province is recognized as a dengue hotspot, with over 45,000 cases reported during the 2014 outbreak [6], and more than 17,000 cases reported in 2024, the most severe outbreak in the past five years [7]. Moreover, the high rate of asymptomatic infections suggests that the true burden of dengue is likely severely underestimated [2]. These trends underscore the persistent threat dengue poses to global public health and highlight its growing economic and healthcare burden [4,8,9].

Dengue transmission is driven by a complex interplay of external factors such as climatic, ecological, and social factors. Temperature directly affects the extrinsic incubation period of the dengue virus [6], while the dynamics of its primary vectors, such as *Aedes aegypti* and *Aedes albopictus*, are sensitive to climate variables such as temperature, precipitation, and humidity [10]. These climatic factors influence mosquito development rates, reproductive cycles, and biting frequency, which in turn affect the intensity and timing of dengue outbreak [11]. Public health interventions also play a critical role [12]; for example, during the COVID-19 pandemic in 2020, dengue cases in Guangdong experienced a sharp decline, demonstrating the substantial impact of non-pharmaceutical interventions (NPIs) on transmission dynamics. Given the recurrent nature of dengue epidemics, it is essential to investigate the mechanistic underpinnings of transmission and assess the relative influence of multiple factors, including climate variability and intervention strategies, on outbreak dynamics. Such insights are vital for enhancing early-warning systems and informing more effective, data-driven public health responses at the regional level.

Dengue transmission dynamics have been extensively studied [3,6,13-33]. First, the mechanistic models, such as the classical Susceptible-Infected-Recovered (SIR) framework, was used to simulate epidemic progression. The Ross-Macdonald model, widely used for vector–host interactions in mosquito-borne diseases like malaria and dengue, has been extended by incorporating factors like climate [6,13-19], *Aedes* mosquito density [6,20], medical resources[21], immunization strategy [22,23], media influence[24], and human movement [3,25] to better understand dengue spread, and to discuss the mitigation and control strategy [26,27]. Second, data-driven models use machine learning techniques such as back-propagation neural networks (BPNN), random forests (RF), support vector machines (SVM), and Long Short-Term Memory (LSTM) networks to predict dengue incidence [28,29]. Third, statistical models like regression model [30], autoregressive integrated moving average (ARIMA)[31], generalized linear models (GLMs) [32], and generalized additive models (GAMs) [33] are useful for analyzing lagged and nonlinear effects of climate variables, though their predictive performance often lags behind that of machine learning models. While mechanistic models offer strong theoretical insight, they often fail to fully integrate high-dimensional data, limiting their ability to capture complex spatiotemporal patterns. In contrast, machine learning models excel at identifying nonlinearities and time-dependent relationships, offering higher predictive accuracy but typically lacking in explaining the underlying transmission mechanisms.

Recent studies have increasingly employed different modeling approaches to enhance the understanding of dengue transmission and its predictions. Li et al. (2019) replaced traditional vector compartments with climate-driven mosquito abundance proxies, linking seasonal dengue risk to environmental conditions using the GAM model, and showing how climate variability impacts mosquito populations and dengue dynamics [6]. More recently, Lu et al. (2025) proposed a hybrid model combining an LSTM neural network with a two-layer differential equation model. By jointly estimating mosquito biting rates through both mechanistic and data-driven components, their model successfully predicting dengue incidence in Selangor, Malaysia, under varying climate conditions. This model achieved accurate forecasts 20 to 60 weeks in advance, highlighting the potential of combining deep learning with mechanistic models for early warning systems [34]. Despite progress in dengue modeling, few studies have examined the combined effects of climate, *Aedes* mosquito density, and public health interventions, and the integration of epidemiological models with deep learning remains underdeveloped.

To address these gaps, we propose a hybrid modeling framework that integrates a classical Susceptible-Infected-Recovered (SIR) model with RNN, leveraging deep learning to capture complex spatiotemporal patterns in high-dimensional data, such as climate, mosquito density, and intervention measures, while retaining the interpretability of mechanistic models. Focusing on Guangdong Province, a dengue hotspot in southern China, we examine transmission dynamics from 2016 to 2024. We first evaluate the performance of multiple RNN architectures in predicting adult *Aede*s mosquito density (MOI) using entomological surveillance data from 2016 to 2022 in ten cities. The best-performing model, LSTM-4V, is then used to estimate mosquito density of Guangdong province for 2016–2024, which is incorporated alongside climate variables and quantified intervention intensity into the coupled SIR model. This hybrid approach enhances forecasting accuracy and interpretability, offering a data-informed basis for targeted public health decision-making in Guangdong and other high-risk regions.

## 2. Materials and Methods

### 2.1 Entomological and Climatic Data Collection

Guangdong Province spans both tropical and subtropical regions, with diverse topography and climate conditions that vary markedly across cities. These environmental differences strongly influence the ecology of *Aedes aegypti* and *Aedes albopictus*, the primary dengue vectors, resulting in substantial spatial heterogeneity in mosquito population density [35]. To monitor *Aedes* populations, two standard entomological indices are employed: the Mosquito Ovitrap Index (MOI), representing the proportion of positive ovitraps among all deployed effective traps in a given area and reflecting adult mosquito density, and the Breteau Index (BI), indicating the percentage of positive larval containers per 100 surveyed households or public premises. According to provincial guidelines, biweekly surveillance is conducted from March to October in dengue-endemic areas. The Guangdong Provincial Health Commission reports MOI and BI values every two weeks and classifies sites into four risk levels, controlled, low, medium, and high [7]. A value exceeding 20 for either index signifies elevated dengue transmission risk.

Figures 1a and 1b illustrate the spatial distribution of average MOI and BI values recorded in mid-April of each year from 2016 to 2022 at high-risk sites across various cities in Guangdong Province. Mid-April typically marks the onset of favorable climatic conditions for mosquito breeding, making it a critical time to capture mosquito distribution patterns and the potential risk of dengue transmission. The data reveal three distinct patterns: (1) cities such as Guangzhou and Maoming exhibit relative high MOI and BI values; (2) cities like Shaoguan, Jiangmen, and Shenzhen maintain relatively low values for both indices; and (3) cities such as Shantou, Zhanjiang, and Qingyuan display a high value for one index but not the other. Based on representativeness and geographical distribution, we selected ten cities—Guangzhou, Shenzhen, Shantou, Shaoguan, Jiangmen, Yangjiang, Zhanjiang, Maoming, Zhaoqing, and Qingyuan, for further investigation into the relationship between local climatic conditions and adult mosquito density.

**Figure 1.**
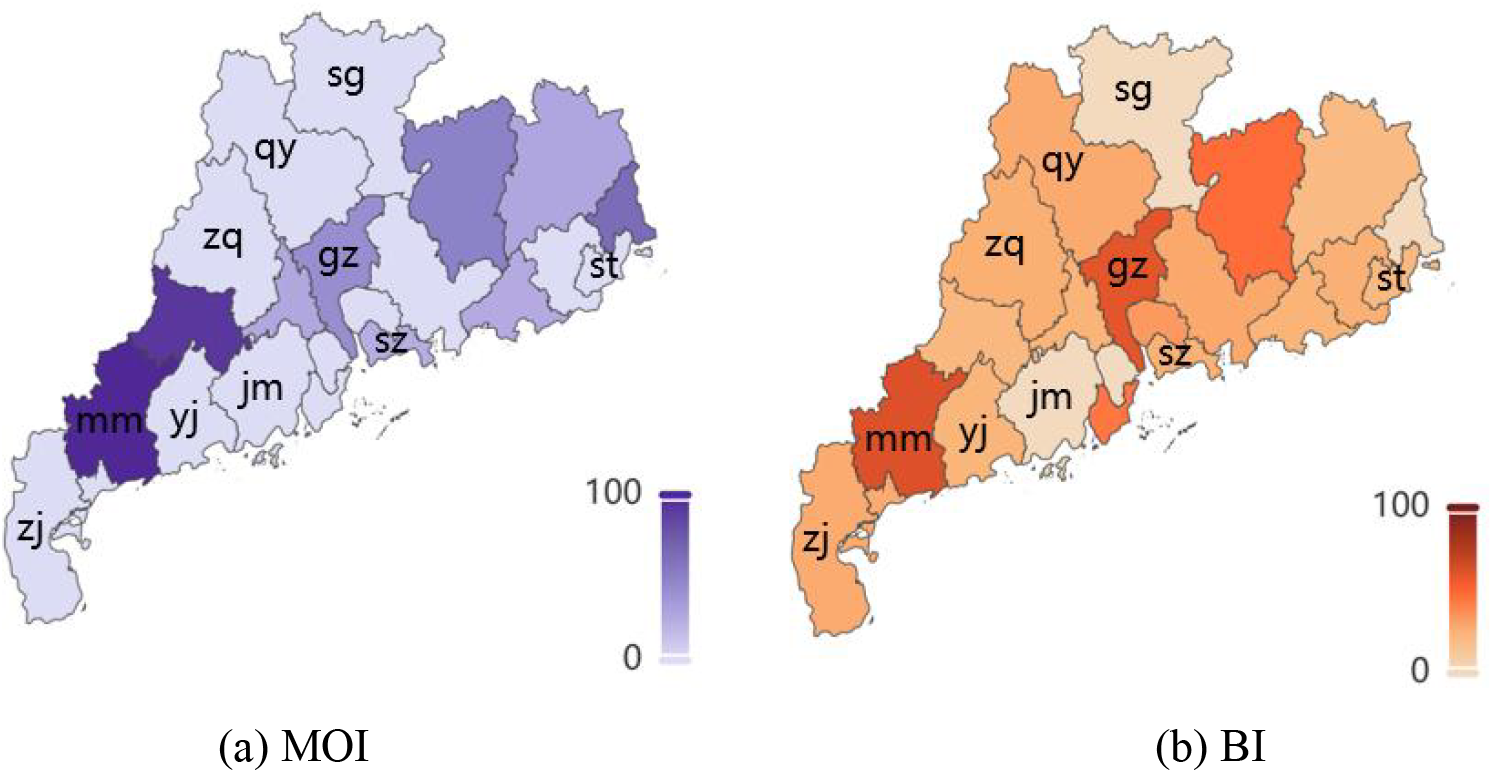
Spatial distribution of MOI and BI across cities in Guangdong Province. The figure illustrates the distribution of the Mosquito Ovitrap Index (MOI) and the Breteau Index (BI) across ten cities of the province: Guangzhou (gz), Shenzhen (sz), Shantou (st), Shaoguan (sg), Jiangmen (jm), Yangjiang (yj), Zhanjiang (zj), Maoming (mm), Zhaoqing (zq), and Qingyuan (qy).

Daily meteorological data for these ten cities, including maximum, minimum, and mean temperature, precipitation, and humidity, were obtained from the National Centers for Environmental Information (NCEI) database maintained by the U.S. National Oceanic and Atmospheric Administration (NOAA) [36]. These data from 2016 to 2022 were used to train and evaluate the performance of neural network models for simulating MOI, and to predict the temporal trends in *Aedes* mosquito density across Guangdong Province from 2016 to 2024.

To account for the recurrent nature of dengue epidemics in Guangdong Province and the potential influence of social distancing measures during the COVID-19 pandemic, we analyzed monthly dengue case counts from 2016 to 2023 and employed the LSTM-SIR model to predict the epidemic trend for 2024. This allowed for a comparative assessment of transmission dynamics before and during the pandemic period. Dengue incidence data were obtained from the Public Health Science Data Center [37]. Population-related parameters used in the compartmental model were derived from the Guangdong Statistical Yearbook 2023, published by Beijing Info Press [38].

### 2.2 Quantifying Intervention Intensity

Following the first confirmed COVID-19 case in Guangdong Province on January 19, 2020, provincial authorities enacted a series of stringent NPIs to contain the outbreak. These included school and workplace closures, suspension of public transportation, stay-at-home mandates, mobility restrictions, and enhanced public health messaging. While primarily aimed at controlling SARS-CoV-2 transmission, these measures also coincided with a sharp decline in reported dengue cases. By restricting human mobility and social interaction, they reduced the frequency of human–mosquito contact and altered patterns of vector exposure, particularly in high-risk urban environments. In addition, strengthened public health messaging likely increased the awareness of vector control practices, such as eliminating standing water or employing personal protective measures.

To evaluate the impact of these interventions on dengue dynamics, we quantified policy intensity using metrics from the Oxford COVID-19 Government Response Tracker (OxCGRT) [39], which standardizes government response indicators across jurisdictions. Policy strength was assessed through the following components: (1) Policy Coverage Scope: A binary indicator (Flag) denotes whether a given policy applied locally (Flag = 0) or across the entire province (Flag = 1). In Guangdong, measures such as school and workplace closures and stay-at-home orders were implemented uniformly province-wide (Flag = 1). (2) Policy Stringency Level: Each policy domain was assigned an ordinal level based on implementation intensity. For example, school closure policies ranged from level 0 (no closure) to level 3 (complete closure of all educational institutions). Policy strictness varied over time, with early phases characterized by stringent lockdowns and later phases involving gradual relaxation. (3) Composite Stringency Index (SI): We used the OxCGRT Stringency Index to represent overall policy intensity. This composite metric integrates nine key intervention categories, including closures, mobility restrictions, and public information campaigns, and normalizes the aggregate value to a 0–1 scale, with higher values reflecting more stringent interventions. A more detailed description of the policy stringency indicator can be found in reference [39].

In our hybrid LSTM-SIR model, the time-varying stringency index *u*(*t*) was incorporated as an exogenous covariate to mechanistically represent public health intervention strength. This allowed us to assess the dynamic influence of control policies on dengue transmission and to estimate their suppressive effect on outbreak magnitude. We specifically assume that these interventions reduce the probability of successful mosquito bites on humans. The detailed formulation of this assumption is described in the following section. As the OxCGRT Stringency Index is only available up to February 2023, we assumed *u*(*t*) = 0 from March 1, 2023 onward, reflecting the transition to routine public health management.

### 2.3 The hybrid modelling framework

To explore how climate variability, mosquito abundance, and public health interventions jointly shape dengue transmission dynamics, we developed a hybrid modeling framework that integrates deep learning with a mechanistic SIR model (Figure 2). Specifically, we employed recurrent neural networks (RNNs), including Long Short-Term Memory (LSTM) and Gated Recurrent Unit (GRU) architectures, to learn temporal patterns between climate variables and *Aedes* mosquito density, using two entomological surveillance indices, MOI and BI. Trained on city-level data from 2016 to 2022, the best-performing RNN model was used to generate province-wide predictions of MOI, serving as a proxy for adult mosquito density across Guangdong. These predictions, along with temperature data and intervention intensity scores derived from the Oxford COVID-19 Government Response Tracker (OxCGRT), were incorporated into a temperature-sensitive SIR model. This coupled framework leverages the predictive strength of deep learning and the explanatory power of mechanistic models, enabling robust simulation of dengue spread and providing a flexible platform to evaluate the impact of climate and control strategies on epidemic dynamics.

**Figure 2.**
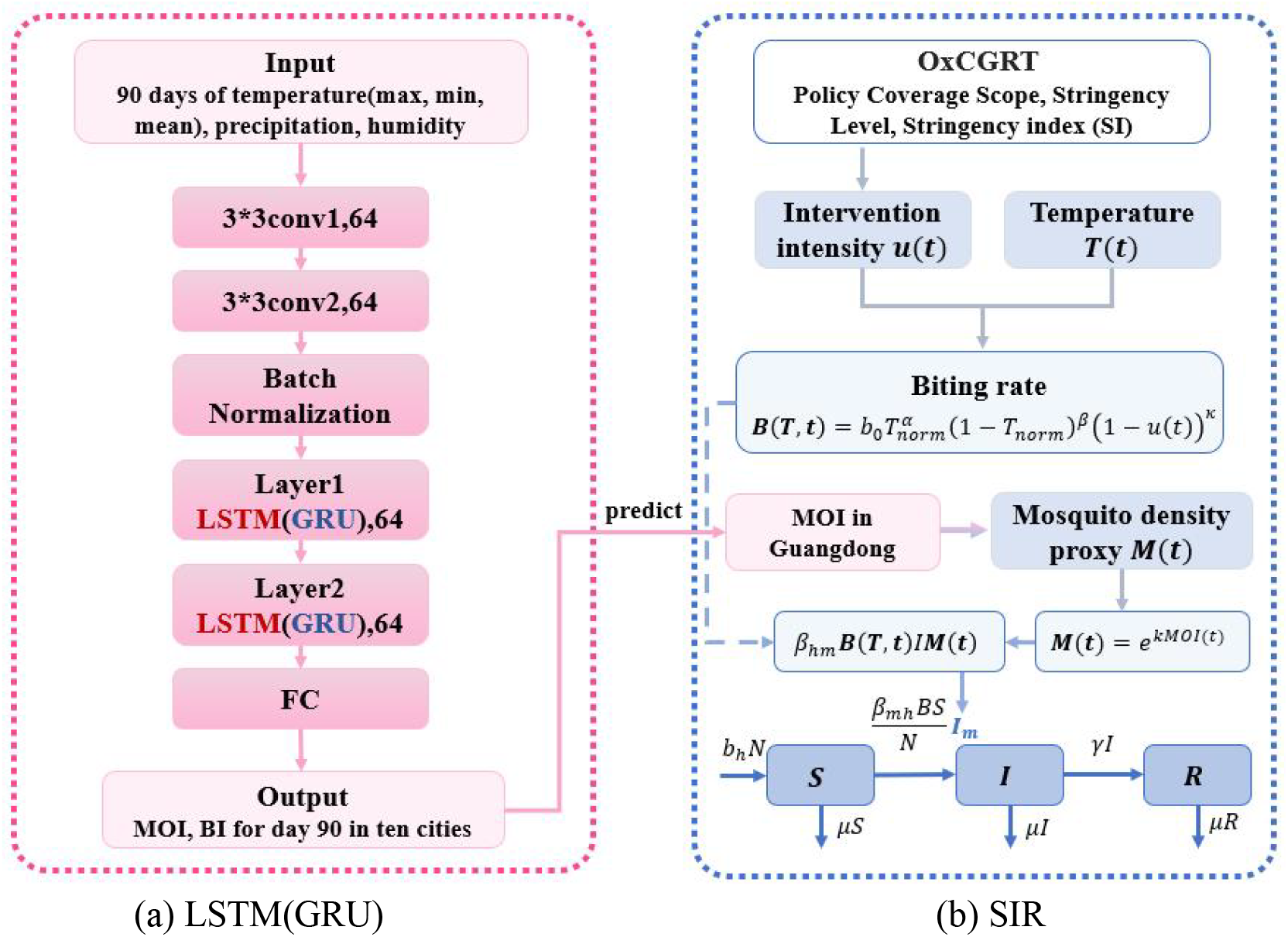
Overview of the hybrid modeling framework integrating deep learning and mechanistic models for dengue transmission in Guangdong Province. Panel (a) illustrates the architecture of the RNN models (LSTM or GRU), (b) depicts an SIR-based mechanistic model to simulate the dengue transmission dynamics, considering climate data and policy intervention strength and mosquito abundance.

#### 2.3.1 Recurrent Neural Networks

To model the relationship between climatic conditions and mosquito density, we employed two RNN architectures, LSTM and GRU, which are well-suited for capturing temporal dependencies in sequential data. Prior studies have demonstrated the feasibility of using neural networks to estimate mosquito abundance directly from climate variables [40]. LSTMs utilize a three-gate structure (input, forget, output) to retain long-range dependencies and mitigate vanishing gradient issues, while GRUs adopt a simplified gating scheme to enhance computational efficiency. Both architectures were implemented and compared in this study.

Given the approximate 30-day lifespan of *Aedes* mosquitoes and the delayed ecological responses to weather, we used a 90-day sequence of daily meteorological inputs[6,40,41], including maximum, minimum, and mean temperature, precipitation, and humidity, to capture both immediate and lagged effects. Due to limited MOI data availability, we modeled MOI and BI jointly to improve stability and prediction accuracy. Figure 2(a) illustrates the architecture of the three models developed: LSTM-4V, GRU-4V, and a variant referred to as LSTM-5V. Each model begins with two 1D convolutional layers (64 filters, kernel size of 3, no padding, stride of 1) activated by ReLU, followed by batch normalization layers to reduce overfitting and enhance generalization. These are followed by two recurrent layers, either LSTM or GRU units, with 64 hidden units each, designed to extract temporal features from the sequential input. A fully connected layer with ReLU activation and *l*_2_ regularization is applied before the final output. All models models generate two outputs corresponding to the predicted MOI and BI values for day 90.

##### Model training

Before training, all meteorological variables were standardized. Temperature values were converted to degrees Celsius, and precipitation data were screened for missing or anomalous values. Missing entries were filled using historical weather records, and outliers were replaced with the mean precipitation from 2016 to 2022. All inputs were then normalized to the [0, 1] range using global minima and maxima, ensuring comparability across features and facilitating more efficient learning of climate–mosquito relationships. Also, we normalized the MOI and BI values for each city using min–max scaling based on global maximum and minimum observations.

To construct a parsimonious and robust predictor set, we implemented a two-step variable selection process. First, a variance inflation factor (VIF) analysis among climate predictors was conducted to formally assess multicollinearity. Predictors with VIF values exceeding 10 were considered to contribute excessive collinearity and were thus candidates for exclusion. Subsequently, we also compared three neural network architectures to further determine the optimal set of climatic inputs for mosquito density prediction. The LSTM-5V model incorporated five variables, daily maximum, minimum, and mean temperature, precipitation, and humidity, while the LSTM-4V and GRU-4V models excluded mean temperature. Each training sample was structured as a pair (*x*_*i*_, *y*_*i*_), where *x*_*i*_ ∈ ℝ^90×4^ for the LSTM-4V and GRU-4V models, representing 90 consecutive days of four climate variables: maximum temperature, minimum temperature, precipitation, and humidity. For the LSTM-5V model, which additionally included mean temperature, *x*_*i*_ ∈ ℝ^90×5^. The corresponding target output *y*_*i*_ ∈ ℝ^1×2^ comprises the MOI and BI values recorded on the 90th day. Samples from the ten selected cities were prepared accordingly and randomly shuffled to eliminate potential spatial–temporal dependencies. The dataset was then split into 80% for training and 20% for validation. The training set was used to optimize the model’s loss function, while the validation set guided hyperparameter tuning (e.g., learning rate) and assessed generalization performance.

The loss function used in this study is defined as follows:

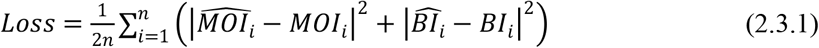

Where 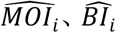 are the predicted values, and *MOI*_*i*_ *BI*_*i*_ are the observed values, and *n* is the total number of samples. To evaluate the models’ overall goodness-of-fit across both outputs, we also computed a joint coefficient of determination (*R*^2^), defined as:

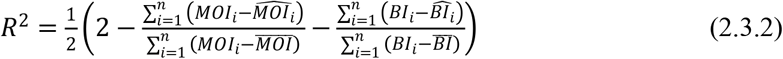

Here,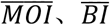 denote the sample means of the observed MOI and BI, respectively.

Model training was performed using the Adam optimizer to minimize the loss function and update parameters. A two-phase training strategy was adopted: an initial phase with a higher learning rate (*α* = 0.01) to accelerate convergence, followed by fine-tuning with a smaller learning rate (*α* = 0.001), which achieved optimal validation performance. To prevent overfitting and retain the best-performing model, early stopping was employed with a patience threshold of 15 epochs. Training was halted if no improvement in loss or *R*^2^ was observed over 15 consecutive validation steps.

##### Model evaluation metrics

To evaluate the accuracy of different models in capturing adult mosquito density, we used the Root Mean Square Error (RMSE) of the predicted MOI as a primary performance metric. RMSE is a widely used indicator that quantifies the deviation between predicted and observed values, providing a balanced measure of prediction bias and variance. The RMSE is computed as:

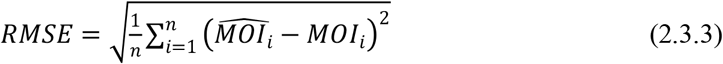

where all variables follow the definitions provided earlier. A lower RMSE indicates better model performance and closer alignment between predicted and actual mosquito density.

#### 2.3.2 Mechanistic model for dengue transmission

To capture the transmission dynamics of dengue fever in Guangdong Province under the combined influence of climate variability, mosquito density, and public health interventions, we developed an extended Susceptible–Infected–Recovered (SIR) compartmental model. The human population was stratified into three epidemiological states: susceptible (*S*), infected (*I*), and recovered (*R*) and the total human population is denoted as *N* = *S* + *I*+ *R*. Additionally, we coupled the SIR framework with LSTM-derived mosquito ovitrap index predictions, *MO* (*t*), yielding a hybrid transmission model (Figure 2), allowing for a more realistic representation of dengue spread influenced by environmental and behavioral drivers.

##### The effect of *Aedes* mosquitoes in the dengue transmission

Dengue virus is transmitted through the *Aedes* mosquitoes, with no direct human-to-human transmission. A susceptible human can become infected following a bite from an infectious mosquito; conversely, a mosquito can acquire the virus after biting an infectious human. The mosquito abundance and its biting behavior plays a pivotal role in shaping the transmission dynamics of the disease.

Given the challenges in directly measuring *Aedes* mosquito abundance, we used the LSTM-predicted MOI as a proxy for active mosquito density, denoted as *M*(*t*). To capture the temporal dynamics of the vector population, we defined mosquito density as an exponential function of MOI: *M* (*t*) = *e*^*kMOI*(*t*)^, where *MOI*(*t*) the time-dependent value predicted by the LSTM-4V model, and *k* is a scaling parameter that quantifies the relationship between MOI and active mosquito density.

To account for the unimodal effect of temperature on mosquito biting activity [41], and the suppressive influence of health interventions, we modeled the per-mosquito biting rate as a time-varying function:

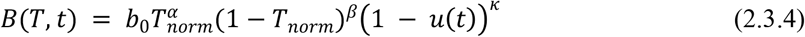

where *T*_*norm*_ = (*T*−*T*_*min*_) / (*T*_*max*_−*T*_*min*_) denotes the daily average temperature normalized by the specified maximum and minimum temperature values, *n*(*t*) represents the normalized intervention intensity (e.g., lockdowns or mobility restrictions), *α* and *β* determine the extent and pattern of temperature’s impact on mosquito activity, and *κ* captures how interventions suppress mosquito biting frequency. Variations in *α* and *β* alter the shape of temperature response curve, resulting in different ranges of mosquito biting frequency. Meanwhile, *κ* modulates the impact of intervention policies: values between 0 and 1 indicate weaker suppression of mosquito biting activity, whereas *κ* > 1 suggests stronger control, leading to a greater reduction in *B*(*T, t*).

##### Mechanistic modeling for dengue transmission

Transmission probabilities between mosquitoes and humans are modeled as time-varying functions influenced by mosquito biting behavior. Specifically, the probability of virus transmission from mosquito to human is given by *β*_*mh*_(*t*) = *β*_*mh*_*B*(*T, t*), and from human to mosquito as *β*_*hm*_(*t*) = *β*_*hm*_*B*(*T, t*), where *B*(*T, t*) is the temperature- and intervention-modulated mosquito biting rate. *β*_*mh*_ and *β*_*hm*_ represent the baseline probabilities of infection per bite from *Aedes* mosquito to human and from human to *Aedes* mosquito, respectively.

The force of infection for susceptible individuals is governed by three factors: (i) the biting and transmission probability from mosquito to human *β*_*mh*_(*t*), (ii) the density of infectious mosquitoes *I*_*m*_, and (iii) the proportion of susceptible individuals in the population (*S*/*N*). The number of infectious mosquitoes is modeled as *β*_*hm*_(*t*)*M*(*t*) *I*, where *M* (*t*) is the active mosquito density, and *I* is the number of infectious individuals. Thus, the rate at which susceptible individuals become infected is *β*_*mh*_ (*t*)*M*(*t*)*β*_*hm*_(*t*)*I S* /*N*.

Infected individuals recover at a rate *γ*, transitioning to the recovered class *R* and losing infectivity.

To account for demographic variability, particularly during the COVID-19 pandemic, we incorporated natural birth and death rates (*b*_*h*_ and *μ*) into the model. Table 2 details all state variables, initial conditions, and parameter definitions. The full system of differential equations is provided below.

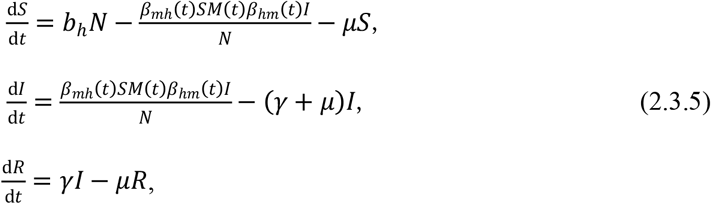

where

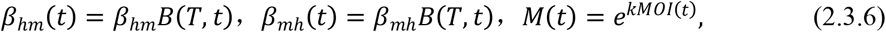

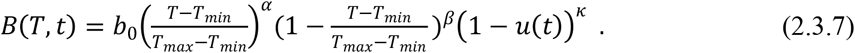

##### Time-varying Reproduction Number

The basic reproduction number *R*_0_ represents the expected number of secondary cases generated by a single infectious individual introduced into a fully susceptible population during their infectious period [42,43]. As a threshold parameter, *R*_0_ < 1 indicates that the infection will eventually die out, whereas *R*_0_ > 1 suggests the potential for sustained transmission and endemicity. Unlike *R*_0_, the time-varying reproduction number *R*_*t*_ captures the real-time transmission potential of the disease by accounting for dynamic changes in climate, vector abundance, and intervention measures at time t. As such, *R*_*t*_ offers a more precise and responsive metric for tracking epidemic trends, evaluating intervention effectiveness, and informing adaptive public health strategies. In this study, we estimate *R*_*t*_ using the next-generation matrix approach applied to the extended SIR framework [43].

At the disease-free equilibrium *x*_0_ = (*N*,0,0)^T^, the new infection matrix ℱ and the transition matrix 𝒱 are defined as:

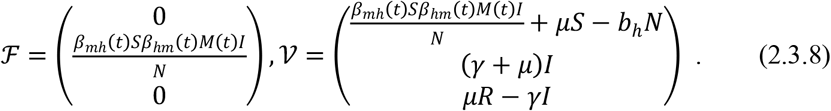

Considering only one infected compartment (*I*), the corresponding scalar components of the matrices are:

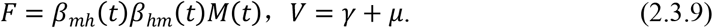

The resulting expression for the time-varying reproduction number is:

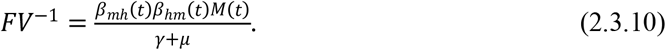

Hence,

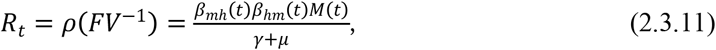

then we have

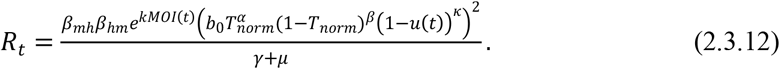

##### Model fitting and parameter estimation

To estimate key parameters in the mechanistic SIR model, we employed a Partially Observed Markov Process (POMP) framework, which enables statistical inference for dynamic systems whose internal states are not directly observable but can be inferred from noisy, incomplete data [44,45]. In our case, the true trajectory of dengue virus transmission is unobservable, and inference is based on monthly reported dengue case counts. We modeled the number of new infections per month using a negative binomial distribution, with the mean equal to the monthly increment in the infectious compartment *I* . To approximate the continuous nature of dengue transmission, we used Euler discretization with a daily time step to simulate system dynamics.

To ensure biological plausibility and facilitate parameter identifiability, we imposed biologically informed constraints: all parameters were required to be non-negative; transmission probabilities *β*_*hm*_ and *β*_*mh*_ were bounded between 0 and 1; the specified maximum and minimum temperature *T*_*max*_ and *T*_*min*_ were respectively set within 13–15°m and 34–36°m; and the mosquito density scaling factor *k* was restricted to the interval [0, 1]. The average time to recovery was fixed at 6 days based on prior studies (see Table 1).

**Table 1.**
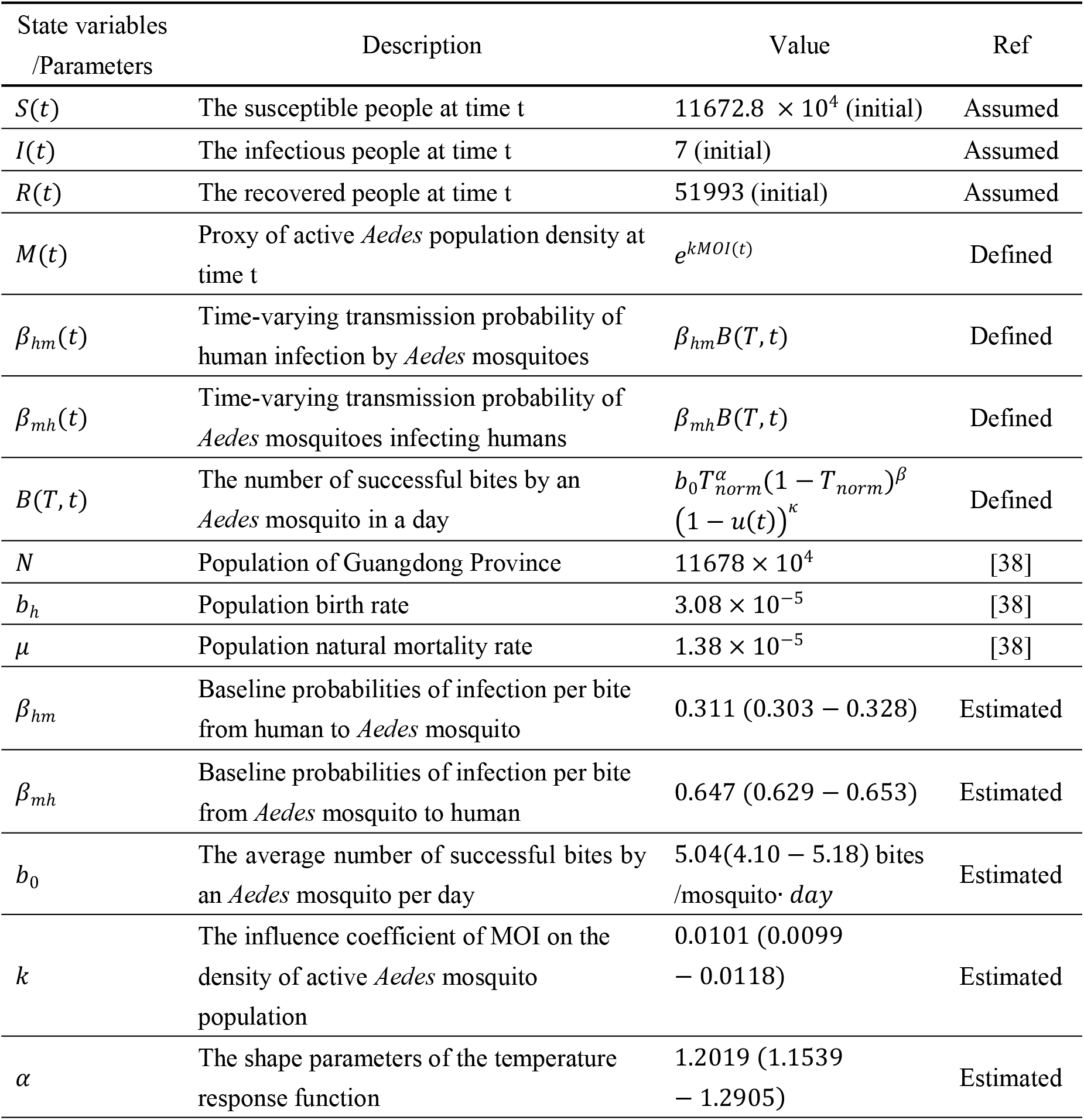
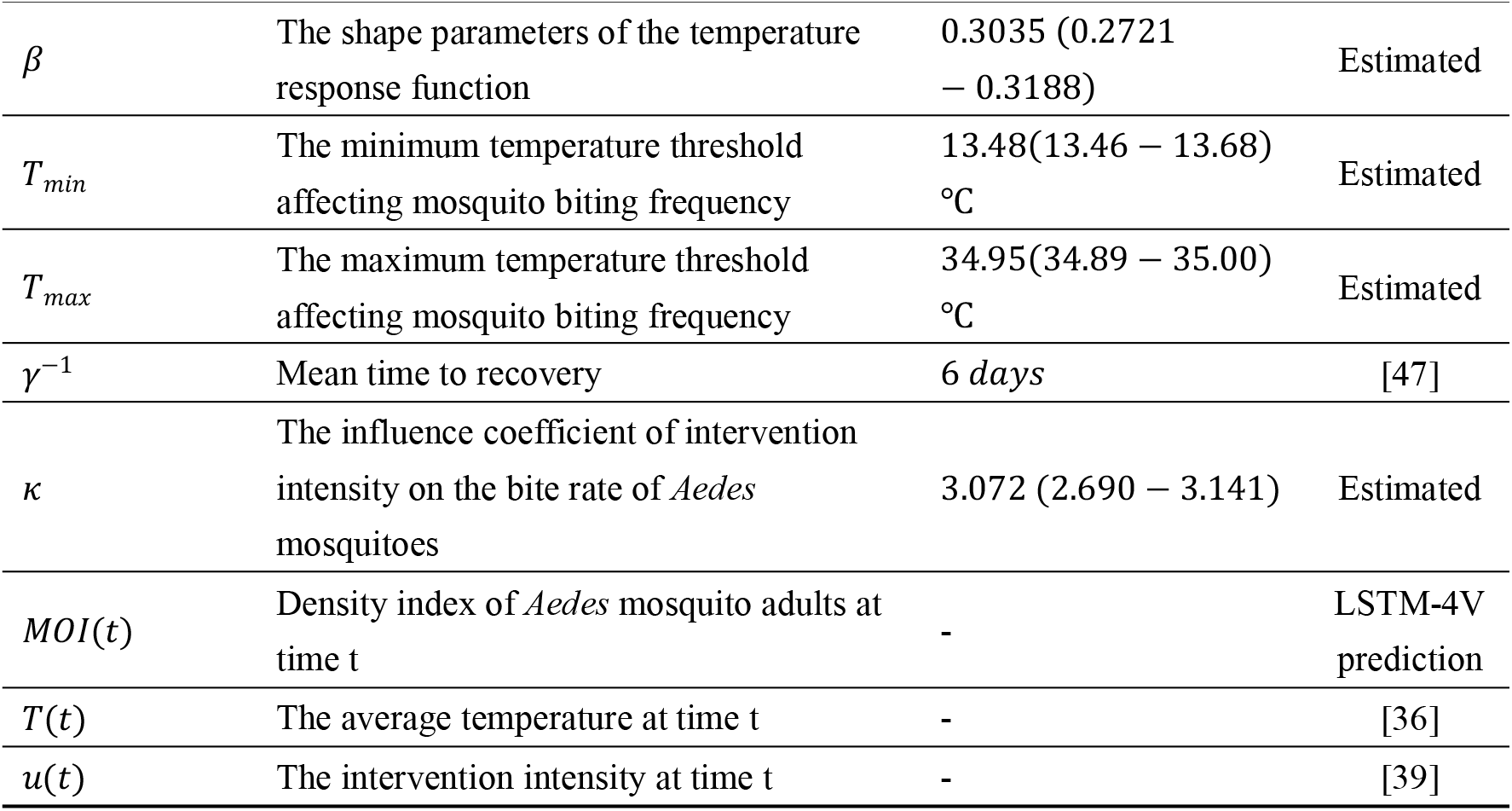
The description and value of state variables, parameters used in the mechanistic model of dengue transmission in Guangdong province.

Parameter estimation was performed using the iterated filtering algorithm [46], with a stochastic perturbation step size of 0.01 for each parameter. This likelihood-based method iteratively maximizes the log-likelihood to obtain optimal parameter values. All possible combination of *T*_*min*_ ∈ {13, 14, 15} and *T*_*max*_ ∈ {34, 35, 36} were evaluated to ensure optimal model performance. The best-fitting LSTM-SIR model yielded a log-likelihood of –458.96, obtained after 1,000 iterations and 20 particles per iteration. The final estimates are reported as median values along with their 95% confidence intervals (see Table 1).

##### Scenario analysis

To evaluate the relative contributions of climatic and policy-driven factors to dengue transmission, we conducted two counterfactual experiments. In the first, all model parameters were held constant while varying the intervention intensity function *u*(*t*) during the period from 2020 to the end of February 2023, to simulate alternative public health responses. Specifically, we modeled two hypothetical public health scenarios by adjusting the values of *u* (*t*) . The first scenario assumed a low intervention level (*u* (*t*) = 0.0556), representing minimal governmental control, e.g., only public health messaging without restrictions on work, school, or transport. Behavioral changes under this scenario were largely voluntary. Protective behavior in this case was primarily voluntary. The second scenario assumed a moderate intervention level (*u*(*t*) = 0.4259), reflecting stronger but non-lockdown measures, such as bans on large gatherings (limited to 11–100 people), enhanced health screening at points of entry, and intensified risk communication via social media.

In the second, we evaluated the effect of temperature on dengue transmission by removing the climate-sensitive term 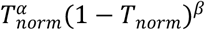 from the mosquito biting rate function *B*(*T, t*) . While keeping mosquito density *M*(*t*) and intervention intensity *n*(*t*) fixed, we re-estimated the model to assess its fit to observed case data in Guangdong. This resulted in a simplified variant of the model, hereafter referred to as **SIR_NoTempBite**, where

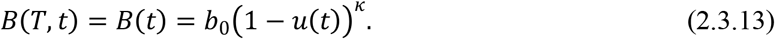

## 3. Results

### Performance comparison of RNN models for mosquito density prediction

We evaluated the predictive performance of three RNN models, LSTM-4V, LSTM-5V, and GRU-4V, for estimating mosquito ovitrap index (MOI) across ten cities in Guangdong Province. The LSTM-5V model contained the highest number of parameters (88,642), followed by LSTM-4V (83,970), and GRU-4V (67,842). Table 2 summarizes the loss values and coefficients of determination (*R*2) for each model on the training and validation sets under the final selected hyperparameters. All models performed well on the training data, with LSTM-5V and GRU-4V achieving *R*2 values exceeding 0.99, and LSTM-4V slightly lower but still above 0.98. These results highlight the capability of RNNs to capture complex temporal dependencies in high-dimensional spatiotemporal data. However, GRU-4V exhibited a noticeable performance drop on the validation set, suggesting limited generalization compared to LSTM-4V and LSTM-5V. While LSTM-5V achieved the best training fit, LSTM-4V demonstrated the most robust validation performance overall.

**Table 2.** The loss value and determination coefficient of different models in the training set and validation set.

| Model | Training sets |  | Validation sets |  |
| --- | --- | --- | --- | --- |
| | *Loss* | $R^2$ | *Loss* | $R^2$ |
| LSTM-4V | 3.1914e-4 | 0.9895 | 0.0036 | 0.8471 |
| LSTM-5V | 1.6075e-4 | 0.9965 | 0.0049 | 0.7883 |
| GRU-4V | 2.0733e-4 | 0.9958 | 0.0072 | 0.6873 |

To further assess model accuracy, we used each trained network to simulate MOI dynamics and computed RMSE for each city (Table 3). Both LSTM-4V and LSTM-5V consistently outperformed GRU-4V, confirming the advantage of LSTM-based architectures in long-term MOI forecasting. While LSTM-5V showed marginally better performance in Guangzhou, Shaoguan, and Zhanjiang, LSTM-4V yielded superior results in the remaining cities and achieved the best overall accuracy. These findings indicate that incorporating average temperature failed to enhance the model predictive performance and instead introduced redundant complexity into the predictor set. Despite the strong overall performance of LSTM-4V, the variability in RMSE across cities reveals constraints in capturing spatial heterogeneity, suggesting the potential need for additional contextual features or recalibration in locally sensitive regions.

**Table 3.**
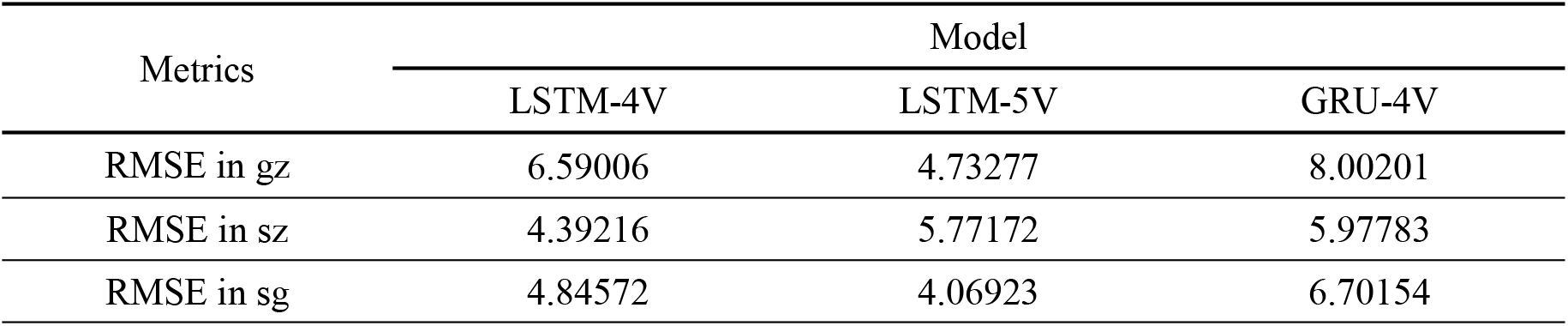
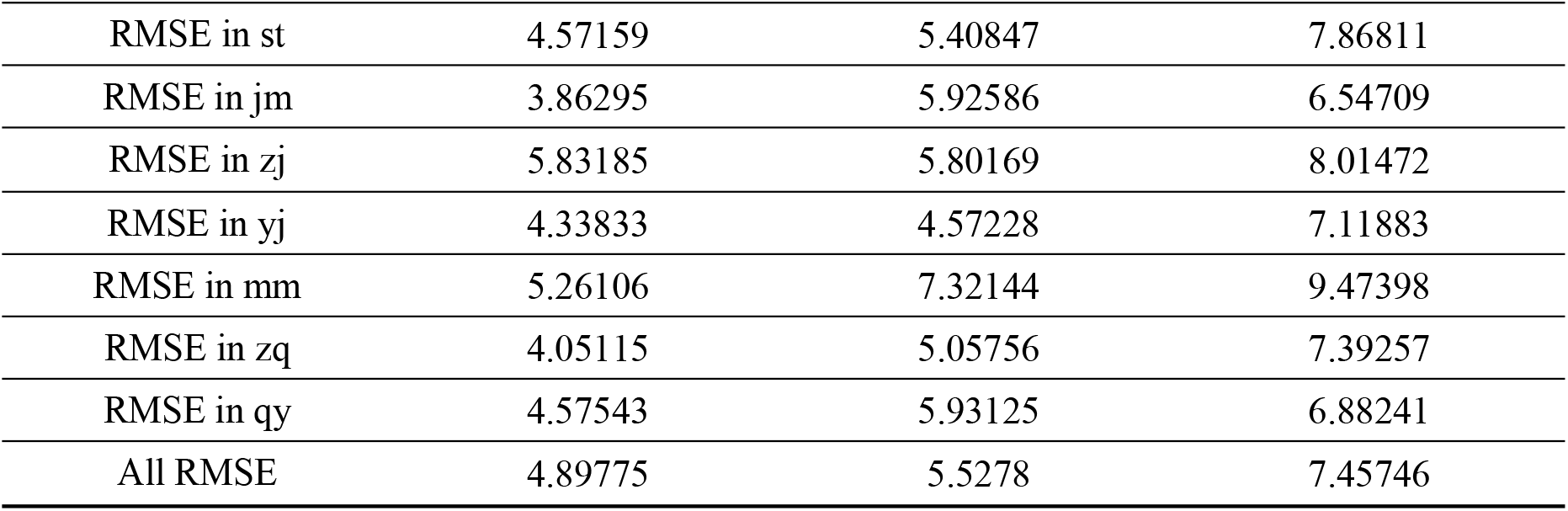
The results of model evaluations.

| Metrics | Model |  |  |
| --- | --- | --- | --- |
|  | LSTM-4V | LSTM-5V | GRU-4V |
| RMSE in gz | 6.59006 | 4.73277 | 8.00201 |
| RMSE in sz | 4.39216 | 5.77172 | 5.97783 |
| RMSE in sg | 4.84572 | 4.06923 | 6.70154 |
|  | LSTM-4V | LSTM-5V | GRU-4V |
| RMSE in st | 4.57159 | 5.40847 | 7.86811 |
| RMSE in jm | 3.86295 | 5.92586 | 6.54709 |
| RMSE in zj | 5.83185 | 5.80169 | 8.01472 |
| RMSE in yj | 4.33833 | 4.57228 | 7.11883 |
| RMSE in mm | 5.26106 | 7.32144 | 9.47398 |
| RMSE in zq | 4.05115 | 5.05756 | 7.39257 |
| RMSE in qy | 4.57543 | 5.93125 | 6.88241 |
| All RMSE | 4.89775 | 5.5278 | 7.45746 |

Figure 3(a) and 3(b) present a heatmap of pairwise correlation coefficients among the five climate variables and a VIF analysis, respectively. Strong correlations (*r* > 0.8) were observed among maximum, minimum, and average temperatures, suggesting redundant information. This redundancy may contribute to accumulated prediction error and reduced generalizability. Consistently, VIF analysis revealed that mean temperature had a VIF value of 13.153, while all other climatic variables had values all below 10, further confirming its pronounced redundancy. Taken together, LSTM-4V provided the most accurate and stable performance across cities, balancing model complexity with predictive capability. Accordingly, although initially included in LSTM-5V model, mean temperature was ultimately excluded to reduce multicollinearity risk, leaving maximum temperature, minimum temperature, precipitation, and humidity as the optimal input feature combination.

**Figure 3.**
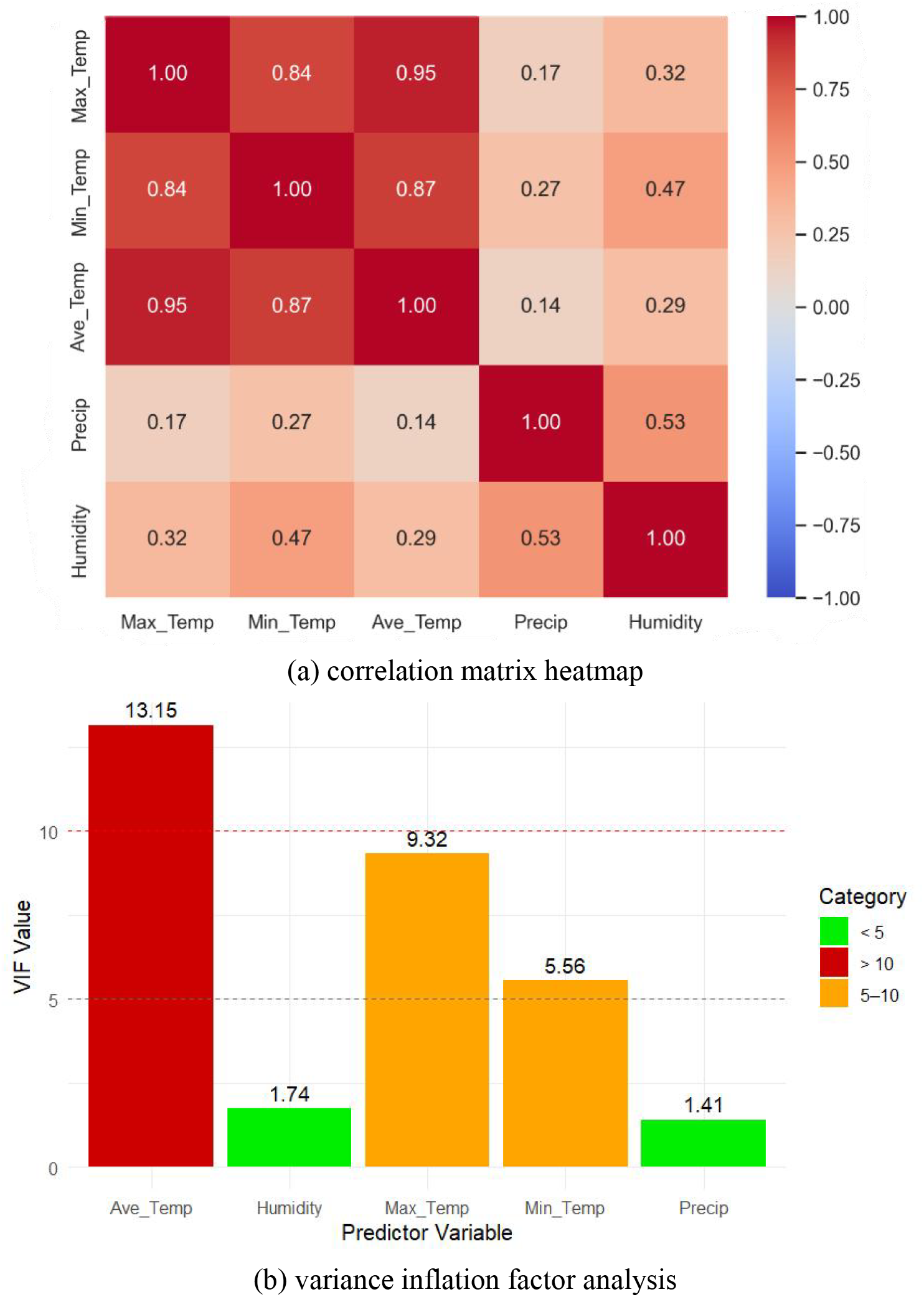
Assessment of correlation and multicollinearity among climate variables.

### Visual validation of LSTM forecasting performance

Figure 4(a) and 4(b) present the prediction results of the LSTM-4V model for Jiangmen and Zhaoqing from 2016 to 2022, two cities where the model achieved the highest evaluation scores. Overall, the LSTM-4V predictions closely align with observed data at most time points, demonstrating the model’s capability to effectively capture the dynamic fluctuations in adult mosquito population density. Nevertheless, localized deviations were observed, potentially attributable to heterogeneity in vector surveillance protocols and mosquito control interventions. Despite such discrepancies, the overall RMSE results confirm that the LSTM-4V model reliably captured the broader temporal trends and maintained robust performance across diverse climatic and geographic conditions.

**Figure 4.**
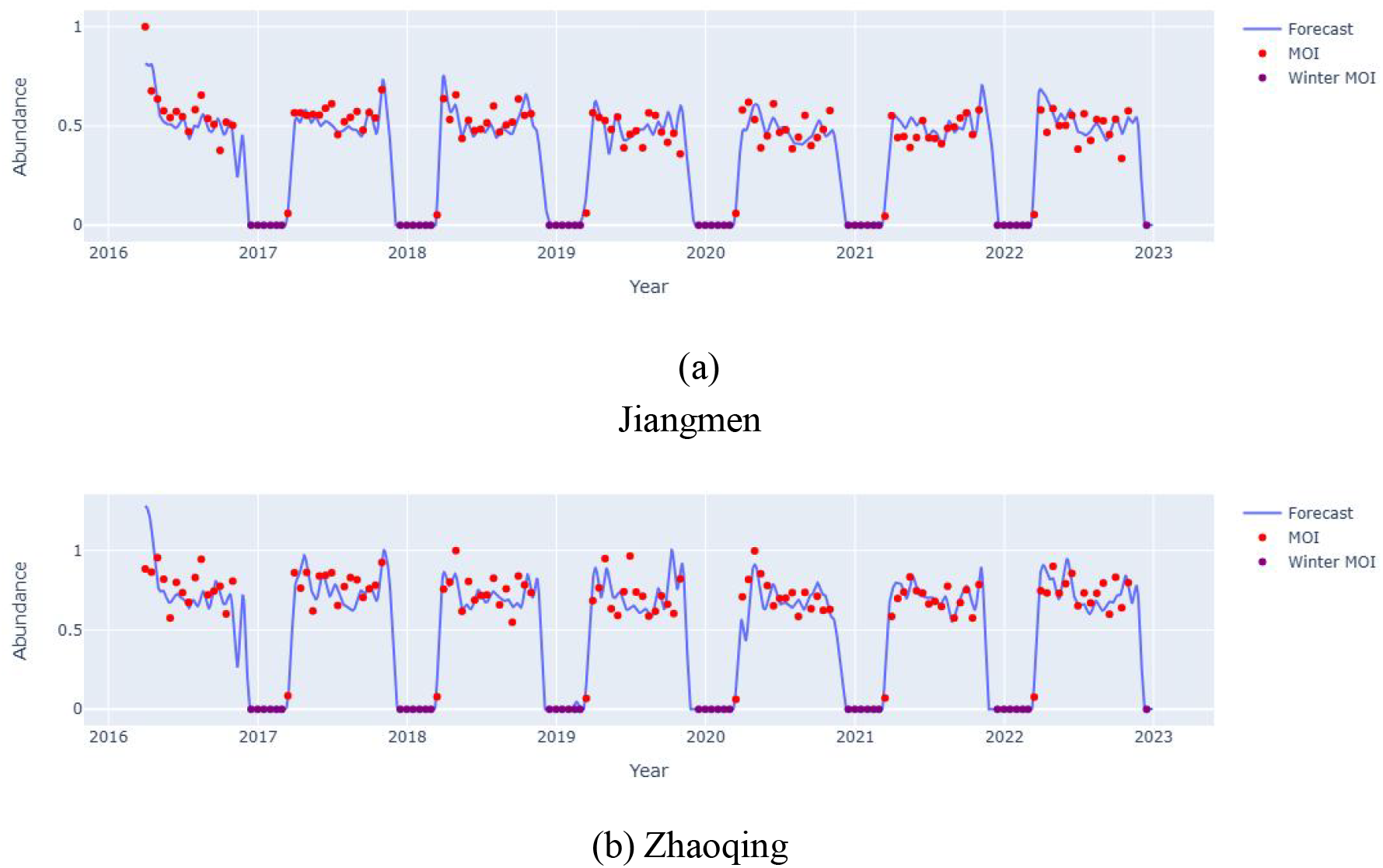
The LSTM-4V model fitting results of MOI. The red dots represent observed MOI values, blue curves denote LSTM-4V predictions, and purple dots indicate periods of mosquito dormancy (MOI set to zero).

Figure 5 shows the LSTM-4V model’s predicted trend of adult *Aedes* mosquito density across Guangdong Province. The results reveal a clear seasonal pattern in mosquito abundance, characterized by periodic fluctuations over time. These predictions provide essential data inputs for subsequent calibration of the SIR transmission model.

**Figure 5.**
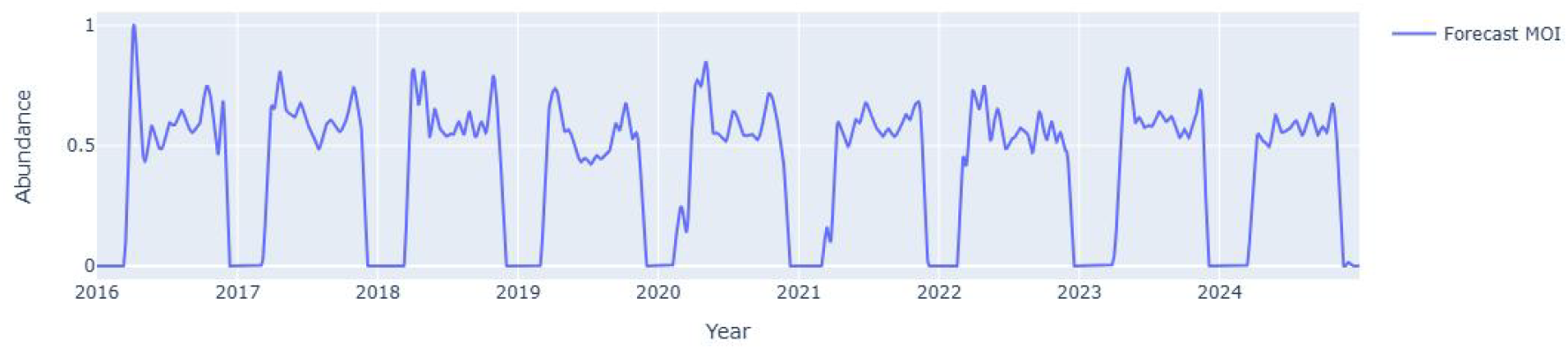
Prediction results of *Aedes* MOI trend in Guangdong Province.

### LSTM-SIR Model fit to observed dengue cases and reproduction number dynamics

Figure 6 illustrates the fit of the LSTM-SIR hybrid model to monthly reported dengue cases in Guangdong Province from 2016 to 2023, along with projections for 2024. The model effectively captures both the timing and magnitude of key outbreaks, including the 2019 epidemic peak and the substantial decline in transmission following the implementation of COVID-19 control measures in 2020. It also successfully reproduces the overall suppression trend during the COVID-19 period and the subsequent rebound in reported cases observed in 2023 as public health restrictions were relaxed. The forecast for 2024 indicates a pronounced resurgence of dengue transmission driven by the continued easing of public health interventions, with the predicted seasonal dynamics closely mirroring pre-pandemic outbreak patterns.

**Figure 6.**
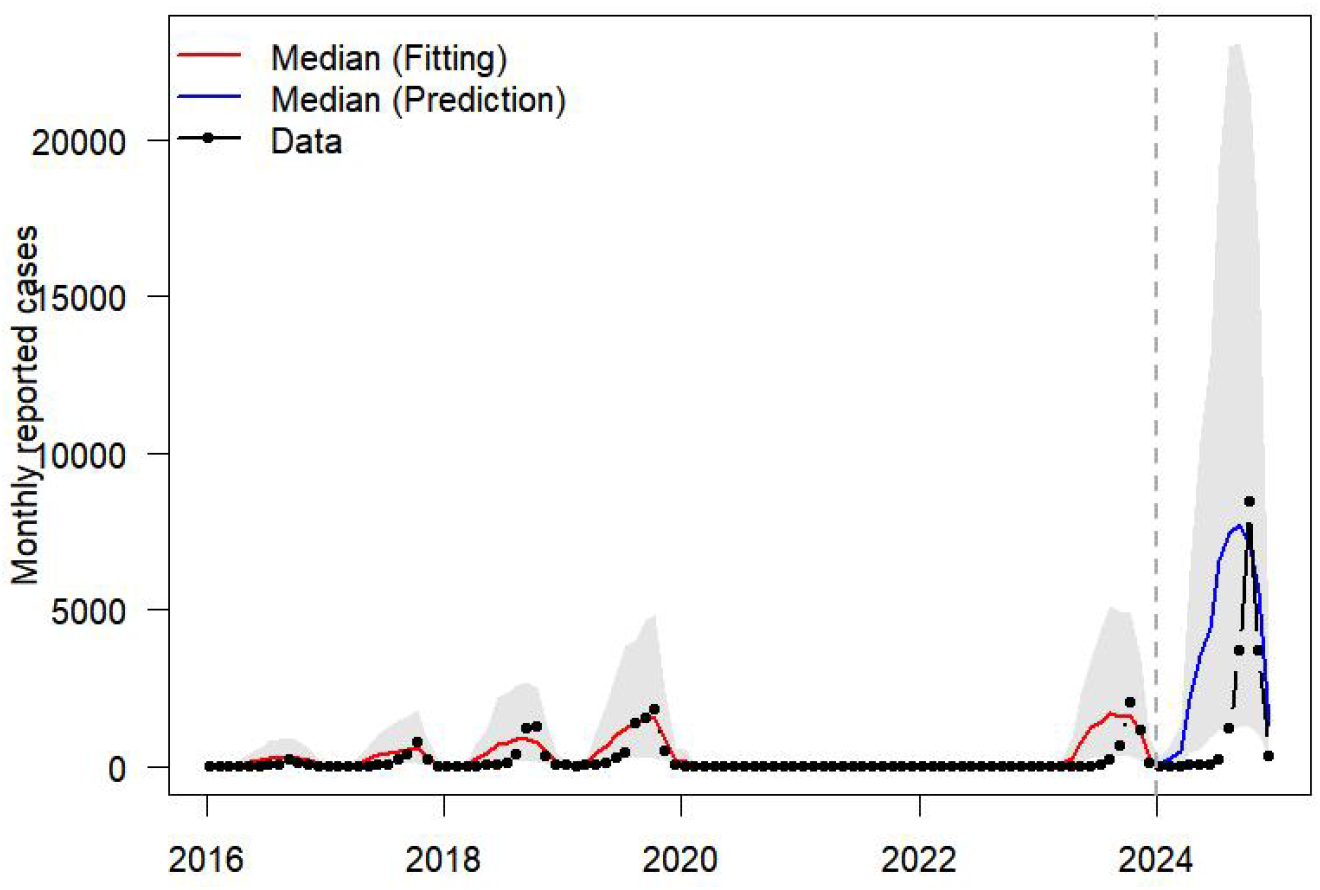
LSTM-SIR model fitting results. The red line represents the median cases of 1,000 simulations, the bule line indicates the median predicted cases of 1000 simulations, the shaded area denotes the 95% confidence interval, and the black dots show observed cases.

Figure 7 depicts the estimated time-varying reproduction number *R*_*t*_, offering mechanistic insights into epidemic dynamics across three distinct periods: the pre-intervention phase (2016–2019), the COVID-19 intervention phase (2020–February 2023), and the post-intervention phase (March 2023–2024). During the baseline period, *R*_*t*_ exhibited a consistent seasonal pattern, rising above 1 in mid-April, peaking around August, and declining below 1 by November, closely tracking the observed seasonal trends in dengue incidence. In contrast, the intervention period was marked by dramatic reductions in dengue incidence, with many months in 2022 reporting zero new cases. These trends coincided with persistently low *R*_*t*_ values, which remained below 1 during the intervention period. Following the end of the COVID-19, the periodic fluctuation of *R*_*t*_ reverted to its pre-intervention baseline pattern, resuming the characteristic rises and falls in line with historical seasonal trends. These results underscore the importance of incorporating behavioral and policy variables into epidemic forecasting models.

**Figure 7.**
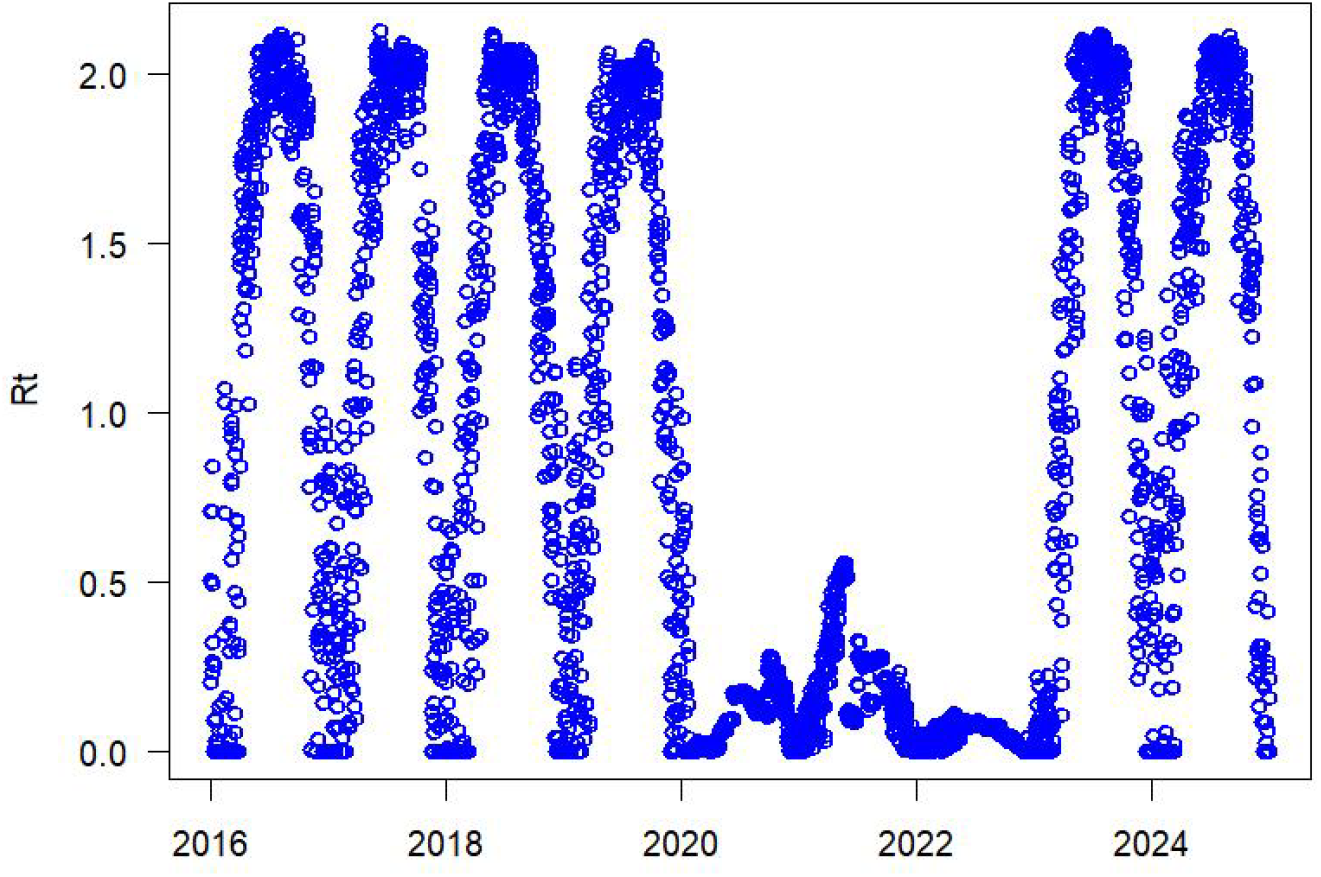
Dengue fever instantaneous reproduction number in Guangdong Province, with blue open circles indicating estimated values.

### Impact of varying policy stringency on dengue transmission

To assess the role of public health interventions in modulating dengue spread, we simulated two scenarios representing differing levels of policy stringency. Figures 8(a) and 8(b) show the projected dengue case counts under weak and moderate intervention intensities, respectively. Under minimal intervention (e.g., public awareness without mobility restrictions), dengue cases surged to over 1,500 at the 2020 peak and subsequently exceeded 3,000 by September–October 2022. Predicted case counts for 2023–2024 persistently surpassed the observed incidence. In contrast, the moderate intervention scenario showed consistently low transmission levels, with monthly incident cases remaining below 100 throughout the period from 2020 to 2022, and falling slightly below the corresponding reported cases from 2023 to 2024. The corresponding time-varying reproduction numbers *R*_*t*_, shown in Figures 8(c) and 8(d), further demonstrate the effectiveness of intervention strategies. Under weak policy enforcement, *R*_*t*_ retained its seasonal pattern and fluctuated between 0 and 2. However, with moderate interventions, *R*_*t*_ remained consistently lower, suppressing transmission and reducing cumulative infections by an estimated 46,120 cases compared to the low-intervention scenario.

**Figure 8.**
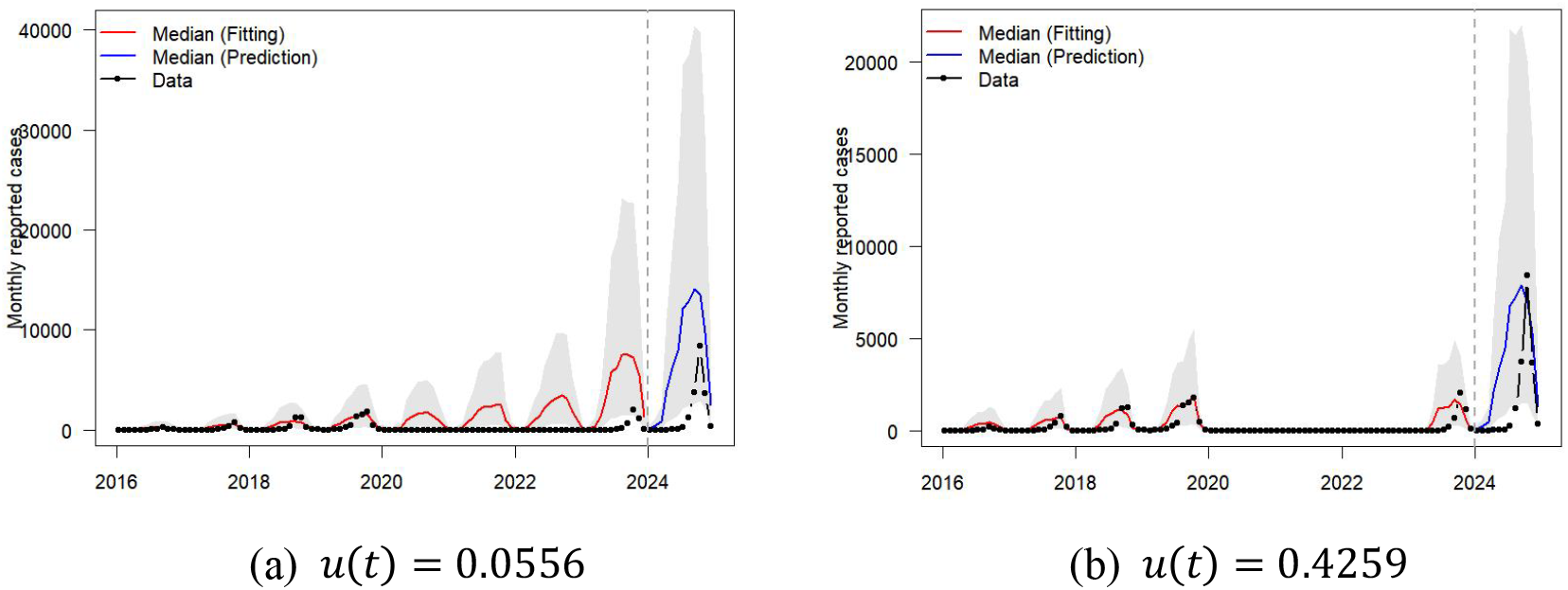
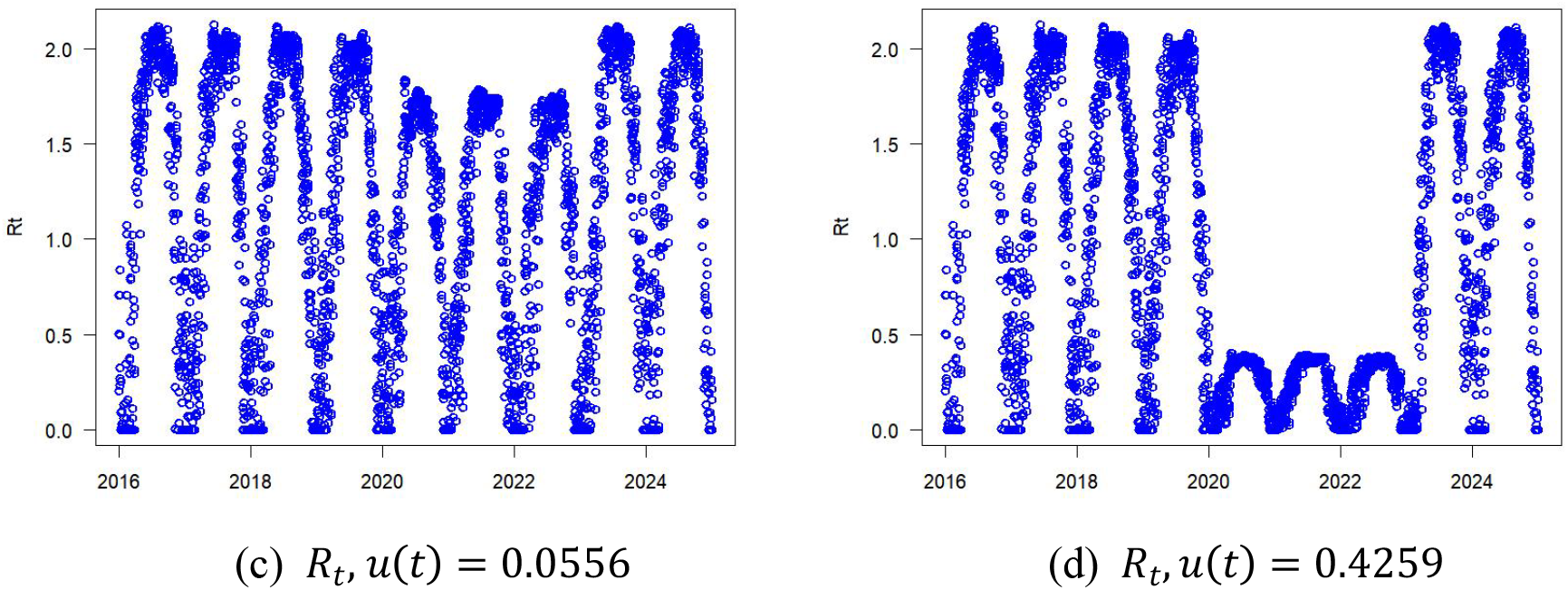
Model fitting results and the time-varying reproduction numbers *R*_*t*_.

These findings suggest that even in the absence of strict lockdowns, moderate public health measures, such as limiting gatherings and promoting risk awareness, can substantially mitigate dengue transmission. Conversely, insufficient intervention may allow the epidemic to follow its typical seasonal trajectory, resulting in significant case burdens.

### Temperature-driven biting behavior is essential for capturing dengue seasonality

To isolate the role of temperature in mosquito biting behavior, we constructed a simplified SIR_NoTempBite model that omits temperature effects from the biting rate formulation. The model was recalibrated using iterated filtering, resulting in a log-likelihood of -635.94. Figure 9 displays the simulated monthly dengue cases across Guangdong Province from 2016 to 2024 under this assumption. The simulation reveals a clear misalignment with observed data. During the low-transmission season (December to March), the model consistently overestimates case numbers, whereas during peak transmission months (September to October), it underestimates cases. Notably, the predicted curve lacks the characteristic seasonal oscillations and instead displays a near-linear increase, failing to reproduce the seasonality of dengue outbreaks.

**Figure 9.**
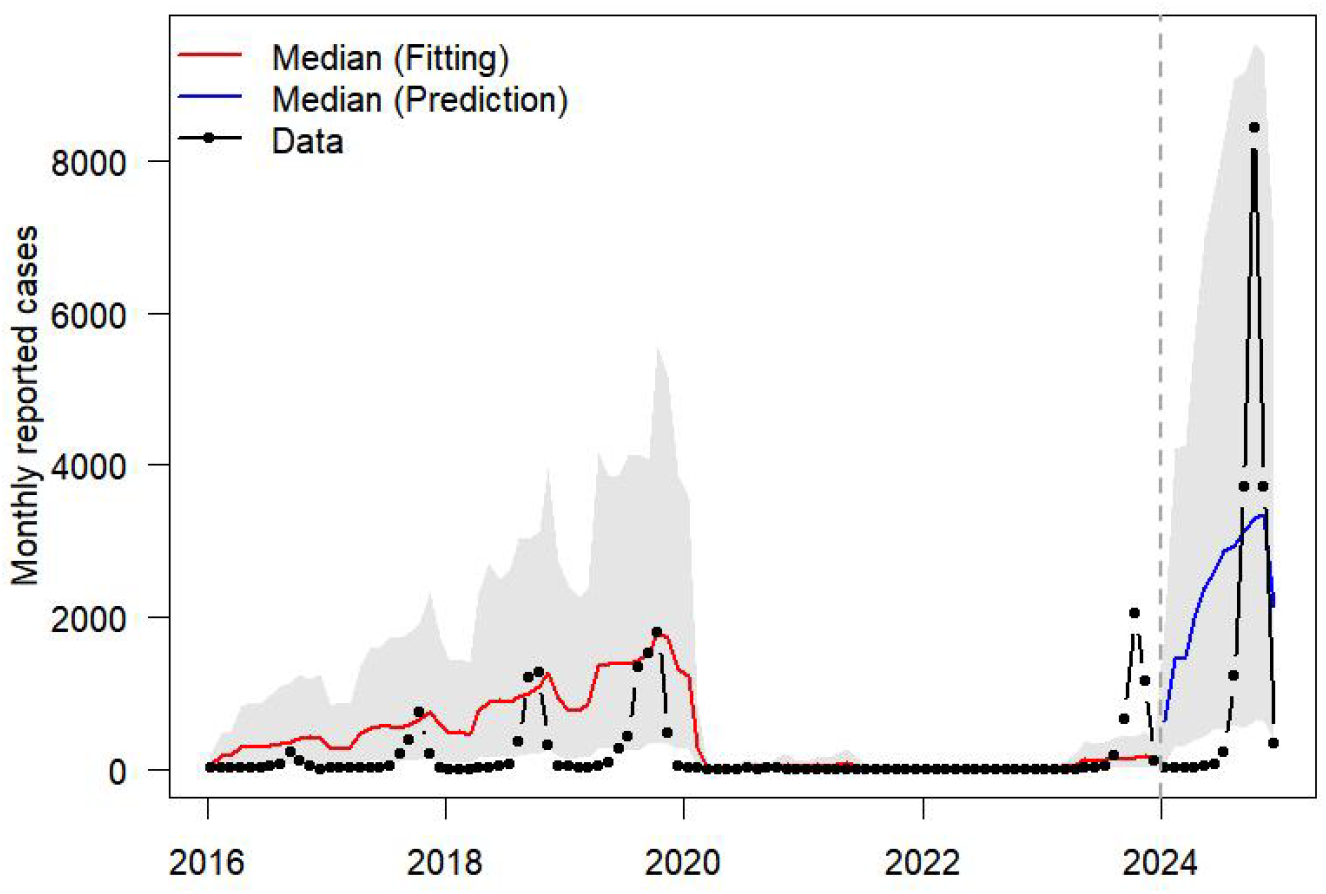
The model simulation results of SIR_NoTempBite.

To quantitatively assess model performance, we compared the SIR_NoTempBite and original SIR models using log-likelihood (LogLik), corrected Akaike Information Criterion (AICc), and mean squared log error (MSLE). As summarized in Table 4, the SIR_NoTempBite model performs worse across all metrics, highlighting the critical role of temperature in accurately capturing dengue dynamics. These findings underscore the essential role of temperature in modulating mosquito biting rates and driving dengue seasonality; omitting this factor significantly reduces model accuracy and limits its forecasting utility for dengue epidemics.

**Table 4.** The comparison of SIR and SIR_NoTempBite model.

| Model | Np | Nmif | Loglik | AICc | MSLE |
| --- | --- | --- | --- | --- | --- |
| SIR | 20 | 1000 | -458.9633 | 948.3656 | 3.221934 |
| SIR_NoTempBite | 20 | 1000 | -635.9444 | 1291.982 | 6.603473 |

## 4. Discussion

In this study, we developed a hybrid modeling framework that couples neural network predictions with a mechanistic compartmental model to investigate the impact of climate, *Aedes* mosquito density, and public health interventions on dengue transmission dynamics in Guangdong Province. We first evaluated the predictive performance of multiple RNN architectures in estimating mosquito density across ten representative cities. The best-performing model (LSTM-4V) was then used to generate province-wide mosquito density estimates, which were subsequently standardized and integrated with climatic variables and policy intervention indices into a climate-informed, mosquito-driven SIR transmission model. This hybrid framework combines the temporal learning capacity of LSTM with the interpretability of mechanistic modeling, enabling more accurate and biologically grounded insights into the drivers of dengue transmission.

Simulation results demonstrate that the LSTM-SIR hybrid model can accurately capture the temporal evolution of dengue outbreaks under varying climatic and policy conditions. The modeled time-varying effective reproduction number *R*_*t*_ effectively replicates the seasonal and periodic patterns of dengue transmission, lending further support to the model’s validity. Moreover, scenario analyses emphasize the critical roles of both climate and public health interventions in shaping outbreak trajectories. Notably, intervention intensity substantially influenced epidemic magnitude, underscoring the effectiveness of NPIs. Although COVID-era strict lockdown policies are no longer feasible, our findings underscore the continued importance of individual-level protective behaviors. In the absence of strict top-down measures, personal measures such as wearing long sleeves, using mosquito repellents, and avoiding outdoor exposure remain key to reducing transmission risk. Given the strong climate sensitivity of mosquito ecology, preemptive vector control efforts should be launched during periods of elevated mosquito density (e.g., April–May and September–October), especially in anticipation of peak transmission seasons (August–October). Looking forward, integrating climate forecasts with data-driven and mechanistic models offers a promising pathway toward real-time dengue risk prediction and early warning systems. The local public health authorities can adopt adaptive policy interventions with meteorological surveillance to enhance the scientific precision and operational responsiveness of outbreak control.

Despite its strengths, this study has several limitations. First, the current model considers only a single dengue serotype and does not account for the interaction among multiple co-circulating serotypes (e.g., DENV-1, DENV-2), which may influence transmission dynamics and cross-immunity patterns [47]. Second, although both *Aedes aegypti* and *Aedes albopictus* are competent dengue vectors [35], our LSTM training did not distinguish between species nor incorporate mosquito control efforts, which can significantly alter vector abundance. Third, the potential for vertical transmission, where dengue-infected mosquitoes emerge from virus-carrying eggs without needing to bite an infected host, was not included in the current modeling framework [48]. These nonlinearities were not explicitly modeled. Additionally, human mobility, an established driver of spatial spread [3,25], along with urban socioeconomic factors, risk-communication intensity, and land-use patterns, could not be incorporated owing to data limitations.

Control measures are of particular public-health interest. While our results show that changes in human behavior prompted by government interventions can markedly curb transmission, the model does not account for vector-control activities and vaccine strategy. We used the OxCGRT Stringency Index as a proxy for policy intensity, but this index does not capture mosquito-specific control efforts such as larvicide spraying, source reduction, or insecticide use. Incorporating direct vector control metrics in future studies would provide a more precise assessment of intervention impacts on mosquito ecology and disease transmission. Furthermore, two vaccines (Dengvaxia and Qdenga) are already licensed [49,50], and a third candidate (TV003) is in phase III trials [51]. Broad vaccine deployment could fundamentally alter dengue epidemiology, which should be considered in the future context.

In the future work, we will enhance both the modeling architecture and epidemiological detail. Transformer-based deep-learning models will be explored to better capture complex temporal dependencies in high-resolution climatic data. Incorporating human-mobility information will allow spatially explicit simulations and finer-scale forecasts of dengue spread. Epidemiological realism will be improved by adding vertical transmission within mosquito populations, modeling interactions among co-circulating serotypes, and explicitly representing vector-control measures and vaccine uptake. Together, these advances will yield a more accurate and interpretable forecasting framework, strengthening public-health preparedness and reducing dengue burden through timely, data-driven interventions.

## Data availability statement

All relevant data are contained within the manuscript.

## Author Contributions

Conceptualization: Pei Yuan, Qiyong Liu, Huaiping Zhu.

Data curation: Pei Yuan, Ning Wang.

Formal analysis: Pei Yuan, Ning Wang.

Software: Pei Yuan, Ning Wang.

Supervision: Huaiping Zhu.

Visualization: Pei Yuan, Ning Wang.

Funding acquisition: Pei Yuan, Huaiping Zhu.

Investigation: Pei Yuan, Ning Wang, Qiyong Liu, Huaiping Zhu.

Writing-original draft: Pei Yuan, Ning Wang, Qiyong Liu, Huaiping Zhu.

Writing-review & editing: Pei Yuan, Ning Wang, Qiyong Liu, Huaiping Zhu.

## Reference

1. Xu L, Stige LC, Chan K-S, Zhou J, Yang J, Sang S, et al. Climate variation drives dengue dynamics. Proceedings of the National Academy of Sciences. 2017;114: 113–118. doi:10.1073/pnas.1618558114

2. World Health Organization. Dengue and severe dengue. [cited 17 May 2025]. Available: https://www.who.int/news-room/fact-sheets/detail/dengue-and-severe-dengue

3. Ni H, Cai X, Ren J, Dai T, Zhou J, Lin J, et al. Epidemiological characteristics and transmission dynamics of dengue fever in China. Nat Commun. 2024;15: 8060. doi:10.1038/s41467-024-52460-w

4. The Lancet editorial. Dengue: the threat to health now and in the future. Lancet. 2024 Jul 27;404(10450):311. doi: 10.1016/S0140-6736(24)01542-3.

5. World Health Organization. Global dengue surveillance. [cited 17 May 2025]. Available: https://worldhealthorg.shinyapps.io/dengue_global/

6. Li R, Xu L, Bjørnstad ON, Liu K, Song T, Chen A, et al. Climate-driven variation in mosquito density predicts the spatiotemporal dynamics of dengue. Proceedings of the National Academy of Sciences. 2019;116: 3624–3629. doi:10.1073/pnas.1806094116

7. Guangdong Provincial Health Commission Portal, Guangdong Provincial Health Commission Official Website. [cited 30 May 2025]. Available: https://wsjkw.gd.gov.cn/

8. Paz-Bailey G, Adams LE, Deen J, Anderson KB, Katzelnick LC. Dengue. The Lancet. 2024;403: 667–682. doi:10.1016/S0140-6736(23)02576-X

9. Shepard D S, Undurraga E A, Halasa Y A, et al. The global economic burden of dengue: a systematic analysis. The Lancet infectious diseases, 2016;16(8): 935–941.

10. Iwamura T, Guzman-Holst A, Murray K A. Accelerating invasion potential of disease vector Aedes aegypti under climate change. Nature communications, 2020; 11(1): 2130.

11. Morin CW, Comrie AC, Ernst K. Climate and Dengue Transmission: Evidence and Implications. Environ Health Perspect. 2013;121: 1264–1272. doi:10.1289/ehp.1306556

12. Chen Y, Li N, Lourenço J, Wang L, Cazelles B, Dong L, et al. Measuring the effects of COVID-19-related disruption on dengue transmission in southeast Asia and Latin America: a statistical modelling study. The Lancet Infectious Diseases. 2022;22: 657–667. doi:10.1016/S1473-3099(22)00025-1

13. Lee H, Kim JE, Lee S, Lee CH. Potential effects of climate change on dengue transmission dynamics in Korea. PLOS ONE. 2018;13: e0199205. doi:10.1371/journal.pone.0199205

14. Cheng Q, Jing Q, Spear RC, Marshall JM, Yang Z, Gong P. Climate and the Timing of Imported Cases as Determinants of the Dengue Outbreak in Guangzhou, 2014: Evidence from a Mathematical Model. PLOS Neglected Tropical Diseases. 2016;10: e0004417. doi:10.1371/journal.pntd.0004417

15. Paul K K, Macadam I, Green D, et al. Dengue transmission risk in a changing climate: Bangladesh is likely to experience a longer dengue fever season in the future. Environmental Research Letters, 2021; 16(11): 114003.

16. Bonnin L, Tran A, Herbreteau V, et al. Predicting the effects of climate change on dengue vector densities in Southeast Asia through process-based modeling. Environmental Health Perspectives, 2022; 130(12): 127002.

17. Hayashi K, Fujimoto M, Nishiura H. Quantifying the future risk of dengue under climate change in Japan. Frontiers in Public Health, 2022; 10: 959312.

18. Saeed A, Ali S, Khan F, et al. Modelling the impact of climate change on dengue outbreaks and future spatiotemporal shift in Pakistan. Environmental Geochemistry and Health, 2023; 45(6): 3489–3505.

19. Wang Y, Zhao S, Wei Y, et al. Impact of climate change on dengue fever epidemics in South and Southeast Asian settings: A modelling study. Infectious Disease Modelling, 2023; 8(3): 645–655.

20. Zhang M, Lin Z, Zhu H. The transmission of dengue virus with Aedes aegypti mosquito in a heterogeneous environment. International Journal of Biomathematics, 2021; 14(05): 2150026.

21. Abdelrazec A, Bélair J, Shan C, et al. Modeling the spread and control of dengue with limited public health resources. Mathematical biosciences, 2016; 271: 136–145.

22. Hladish T J, Pearson C A B, Toh K B, et al. Designing effective control of dengue with combined interventions. Proceedings of the National Academy of Sciences, 2020; 117(6): 3319–3325.

23. Xue L, Jin X, Zhu H. Assessing the impact of serostatus-dependent immunization on mitigating the spread of dengue virus. Journal of Mathematical Biology, 2023; 87(1): 5.

24. Aldila D, Ndii MZ, Anggriani N, Windarto Tasman H, Handari BD. Impact of social awareness, case detection, and hospital capacity on dengue eradication in Jakarta: A mathematical model approach. Alexandria Engineering Journal. 2023;64: 691–707. doi:10.1016/j.aej.2022.11.032

25. Harish V, Colón-González F J, Moreira F R R, et al. Human movement and environmental barriers shape the emergence of dengue. Nature Communications, 2024; 15(1): 4205.

26. Xue L, Ren X, Magpantay F, et al. Optimal control of mitigation strategies for dengue virus transmission. Bulletin of Mathematical Biology, 2021; 83: 1–28.

27. Zhang X, Liu X, Li Y, et al. Modelling the effects of Wolbachia-carrying male augmentation and mating competition on the control of dengue fever. Journal of Dynamics and Differential Equations, 2025; 37(1): 149–189.

28. Ren H, Wu W, Li Q, Lu L. Prediction of dengue fever based on back propagation neural network model in Guangdong, China. Chinese Journal of Vector Biology and Control. 2018;29: 221–225. doi:10.11853/j.issn.1003.8280.2018.03.001 (In Chinese)

29. Doni A, Sasipraba T. LSTM-RNN Based Approach for Prediction of Dengue Cases in India. ISI. 2020;25: 327–335. doi:10.18280/isi.250306

30. Hii Y L, Zhu H, Ng N, et al. Forecast of dengue incidence using temperature and rainfall. PLoS neglected tropical diseases, 2012; 6(11): e1908.

31. Thiruchelvam L, Dass SC, Asirvadam VS, Daud H, Gill BS. Determine neighboring region spatial effect on dengue cases using ensemble ARIMA models. Sci Rep. 2021;11: 5873. doi:10.1038/s41598-021-84176-y

32. Abdulsalam FI, Yimthiang S, La-Up A, Ditthakit P, Cheewinsiriwat P, Jawjit W. Association between climate variables and dengue incidence in Nakhon Si Thammarat Province, Thailand. Geospatial Health. 2021;16. doi:10.4081/gh.2021.1012

33. Chen P, Fu X, Ma S, Xu H-Y, Zhang W, Xiao G, et al. Early dengue outbreak detection modeling based on dengue incidences in Singapore during 2012 to 2017. Statistics in Medicine. 2020;39: 2101–2114. doi:10.1002/sim.8535

34. Lu X, Teh SY, Tay CJ, Abu Kassim NF, Fam PS, Soewono E. Application of multiple linear regression model and long short-term memory with compartmental model to forecast dengue cases in Selangor, Malaysia based on climate variables. Infectious Disease Modelling. 2025;10: 240–256. doi:10.1016/j.idm.2024.10.007

35. Li Y, An Q, Sun Z, Gao X, Wang H. Distribution areas and monthly dynamic distribution changes of three Aedes species in China: Aedes aegypti, Aedes albopictus and Aedes vexans. Parasites Vectors. 2023;16: 297. doi:10.1186/s13071-023-05924-9

36. National Centers for Environmental Information (NCEI). [cited 6 Apr 2025]. Available: https://www.ncei.noaa.gov/

37. Public Health Sciences Data Center. [cited 30 May 2025]. Available: https://www.phsciencedata.cn/Share/

38. Beijing Digital Publishing House. Guangdong Statistical Yearbook 2023. 2023. Available: http://tjnj.gdstats.gov.cn:8080/tjnj/2023/

39. Hale T, Angrist N, Goldszmidt R, Kira B, Petherick A, Phillips T, et al. A global panel database of pandemic policies (Oxford COVID-19 Government Response Tracker). Nature Human Behaviour. 2021;5: 529–538. doi:10.1038/s41562-021-01079-8

40. Kinney AC, Current S, Lega J. Aedes-AI: Neural network models of mosquito abundance. PLOS Computational Biology. 2021;17: e1009467. doi:10.1371/journal.pcbi.1009467

41. Costanzo K, Occhino D. Effects of Temperature on Blood Feeding and Activity Levels in the Tiger Mosquito, Aedes albopictus. Insects. 2023;14: 752. doi:10.3390/insects14090752

42. Diekmann O, Heesterbeek JAP, Metz JAJ. On the definition and the computation of the basic reproduction ratio R0 in models for infectious diseases in heterogeneous populations. Journal of Mathematical Biology. 1990;28: 365–382. doi:10.1007/BF00178324

43. Van Den Driessche P, Watmough J. Reproduction numbers and sub-threshold endemic equilibria for compartmental models of disease transmission. Mathematical Biosciences. 2002;180: 29–48. doi:10.1016/S0025-5564(02)00108-6

44. He D, Ionides EL, King AA. Plug-and-play inference for disease dynamics: measles in large and small populations as a case study. Journal of The Royal Society Interface. 2010 [cited 1 Apr 2025]. doi:10.1098/rsif.2009.0151

45. He D, Lin L, Artzy-Randrup Y, et al. Resolving the enigma of Iquitos and Manaus: A modeling analysis of multiple COVID-19 epidemic waves in two Amazonian cities. Proceedings of the National Academy of Sciences, 2023;120(10): e2211422120.

46. Ionides EL, Bretó C, King AA. Inference for nonlinear dynamical systems. Proceedings of the National Academy of Sciences. 2006;103: 18438–18443. doi:10.1073/pnas.0603181103

47. Halstead SB. Dengue. The Lancet. 2007;370: 1644–1652. doi:10.1016/S0140-6736(07)61687-0

48. Zou L, Chen J, Feng X, Ruan S. Analysis of a Dengue Model with Vertical Transmission and Application to the 2014 Dengue Outbreak in Guangdong Province, China. Bull Math Biol. 2018;80: 2633–2651. doi:10.1007/s11538-018-0480-9

49. WHO. Dengue vaccine: WHO position paper, July 2016recommendations. Vaccine 2017; 35: 1200–01

50. European Medicines Agency. Dengue tetravalent vaccine (live, attenuated) Takeda: opinion on medicine for use outside EU. https://www.ema.europa.eu/en/opinion-medicine-use-outside-EU/human/dengue-tetravalent-vaccine-live-attenuated-takeda (accessed Jun 12, 2025).

51. Biswal S, Reynales H, Saez-Llorens X, et al. Efficacy of a tetravalent dengue vaccine in healthy children and adolescents. New England Journal of Medicine, 2019; 381(21): 2009–2019.

